# Gene prioritization based on systems biology revealed new insight into genetic basis and pathophysiology underlying schizophrenia

**DOI:** 10.1101/2020.06.26.20140541

**Authors:** Jia-Feng Li, Lei Wang, Xiao Dang, Wei-Min Feng, Zi-Wei Wang, Yu-Ting Ma, Si-Jie He, Liang Liang, Huan-Ming Yang, Han-Kui Liu, Jian-Guo Zhang

## Abstract

Sequencing-based studies have recognized hundreds of genetic variants that increase the risk of schizophrenia (SCZ), but only a few percents of heritability can be attributed to these loci. It is challenging to discover the full spectrum of schizophrenia genes and reveal the dysregulated functions underlying the disease. Here, we proposed a holistic model for predicting disease genes (HMPDG), a novel machine learning prediction strategy integrated by Protein-Protein Interaction Network (PPIN), pathogenicity score, and RNA expression data. Applying HMPDG, 1946 potential risk genes (PRGs) as a complement of the genetic basis of SCZ were predicted. Among these, the first decile genes were highlighted as high confidence genes (HCGs). PRGs were validated by multiple independent studies of schizophrenia, including genome-wide association studies (GWASs), gene expression studies, and epigenetic studies. Remarkably, the strategy revealed causal genes of schizophrenia in GWAS loci and regions of copy number variant (CNV), providing a new insight to identify key genes in disease-related loci with multi genes. Leveraging our predictions, we depict the spatiotemporal expression pattern and functional groups of schizophrenia risk genes, which can help us figure out the pathophysiology of schizophrenia and facilitate the discovery of biomarkers. Taken together, our strategy will advance the understanding of schizophrenia genetic basis and the development of diagnosis and therapeutics.

## Introduction

Schizophrenia (SCZ) is a severe psychiatric disorder with high heritability estimated around 80%^1^. Currently, sequencing-based studies have found that schizophrenia is driven by different types of genetic variants, which contains hundreds of common variations, de novo mutations, and structural variants^2^. In particular, landmark genome-wide association study (GWAS) of SCZ lead by Psychiatric Genomics Consortium (PGC) has reported 108 genomic loci associations^3^. However, with the limitation of the sample size and analysis methods, the genetic loci previously reported can explain only a fraction of the heritability for schizophrenia ^4^.To unravel the genetic basis of schizophrenia, we need to explore a new strategy to provide a full-scale list for SCZ risk genes.

Recently, at least 20 gene-specific metrics have been developed, which mainly focus on single-gene assessment for Mendelian disease. Yet, the genetic heterogeneity of complex disorders requires us to evaluate disease-associated genes systematically. The biological network provides effective summaries of cellular function by integrating various interaction information and thus the molecular basis of diseases can be enlightened through the biological network, which in turn can figure out the genetic causes behind the diseases. Prior network-based prediction approaches that integrated multi-level information have shown great potential in discovering disease genes. These approaches, however, ignored the characteristics of the gene itself, such as loss-of-function tolerance, evolutionary conservation, which is essential for the assessment of gene pathogenicity. Besides, the network-based method always concentrated on only one side of the characteristics of the network, which are based either on the graph embedding to represent the ‘position’ of every node in the graph or on the topological parameters to assess the importance of nodes^5^.

To meet these challenges, we developed an integrative strategy based on systems biology named holistic model for predicting disease genes (HMPDG). It is an innovative attempt to offer a comprehensive prediction of SCZ-related genes by integrating three layers of information: 1) exhaustive evaluation for each gene in the protein-protein network (PPIN) based on network embedding and topological parameters, 2) assessment of pathogenicity of genes, and 3) the gene-expression level in developing brain. Notably, to the best of our knowledge, it is the first time that combining the biological network characteristic and gene pathogenicity score to provide a full-scale complement for risk genes of SCZ. Especially, the characteristics of the network were obtained by graph embedding and topological parameter assessment, which can preserve not only the network structure but also the centrality information. Applying HMPDG, we provided a full-scale risk gene list of SCZ and, many of these candidate genes have been validated by multiple sequencing studies. We performed systematic analyses on the prediction results, which provided new insights into the spatiotemporal expression pattern of SCZ and functional modules potentially dysregulated in the brain of patients with SCZ. The risk gene set we provided can be a complement of SCZ-related genes and advance the understanding of the schizophrenia mechanism, which is needed to optimize genetic diagnosis and improve treatments. Further, we emphasize that our strategy is widely available in other complex disease studies and will facilitate gene discovery and provide insights into disease.

## Results

### Prediction of schizophrenia risk genes

We constructed an Integrative predict model for predict SCZ-related genes (Fig.1), which obtain an accuracy of 87.5%, and the receiver operating characteristic (ROC) curve showed an area under the curve (AUC) of 0.896. Our strategy showed better performance than existing methods (Supplementary 1). We applied this method to assess all genes from reliable PPIN and obtained 1946 potential risk genes (PRG) for SCZ.

**Figure 1:**
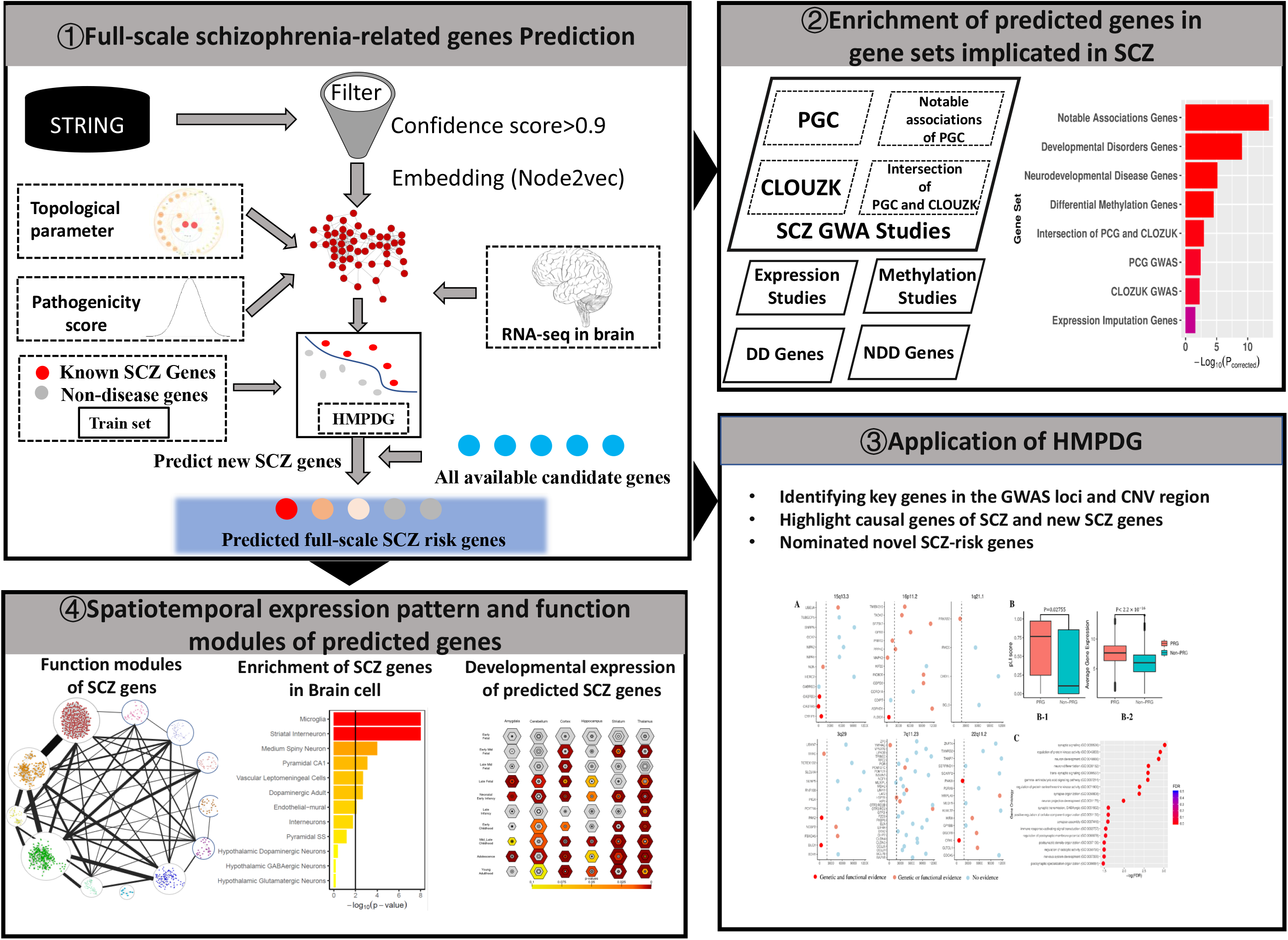
A schematic illustration of the HMPDG framework. (1) Firstly, integrated features from embedding and topological parameters of the network, RNA expression data, and gene pathogenicity score to construct a random forest classifier(HMPDG). Applying HMPDG, the potential SCZ risk genes were predicted.(2) Secondly, we verified the validity of our predicted genes in multiple genetic studies of schizophrenia. Then, (3) we explored the application of the HMPDG to identify causal genes and nominate novel risk genes (4) Finally, we performed systematic analyses to explore the genetic basis and pathogenesis of schizophrenia.

In order to provide additional evidence for predicted accuracy, we executed a systematic empirical evaluation based on the well-established SCZ-related gene set from recent genetic studies. First, we focused on the SCZ-associated genes identified by the GWAS^3,6^. The PRGs show significant enrichment in PGC SCZ-associated gene set (P _corrected_=3.75 × 10^−3^), CLOZUK SCZ-associated gene set (P _corrected_=5.59 × 10^−3^), and the intersection of PGC and CLOZUK(P _corrected_=1.13 × 10^−3^). Second, we turn to the notably associated gene set from PGC^3^ and observed a more significant result (P _corrected_=3.76 × 10^−13^). Third, we evaluated the enrichment of PRGs in the gene sets of expression imputation study and methylation studies of SCZ. As expected, we observed significant enrichment in both expression imputation gene set (P _corrected_=0.026) and differential methylation gene set (P _corrected_=3.03 × 10^−5^)^7-13^. Fourth, we found significant enrichment in the gene sets of developmental disorders (DD, P _corrected_=8.81 × 10^−10^) and neurodevelopmental disorders (NDD, P _corrected_=7.79 × 10^−6^)^14,15^(Fig.2 A).

**Figure 2:**
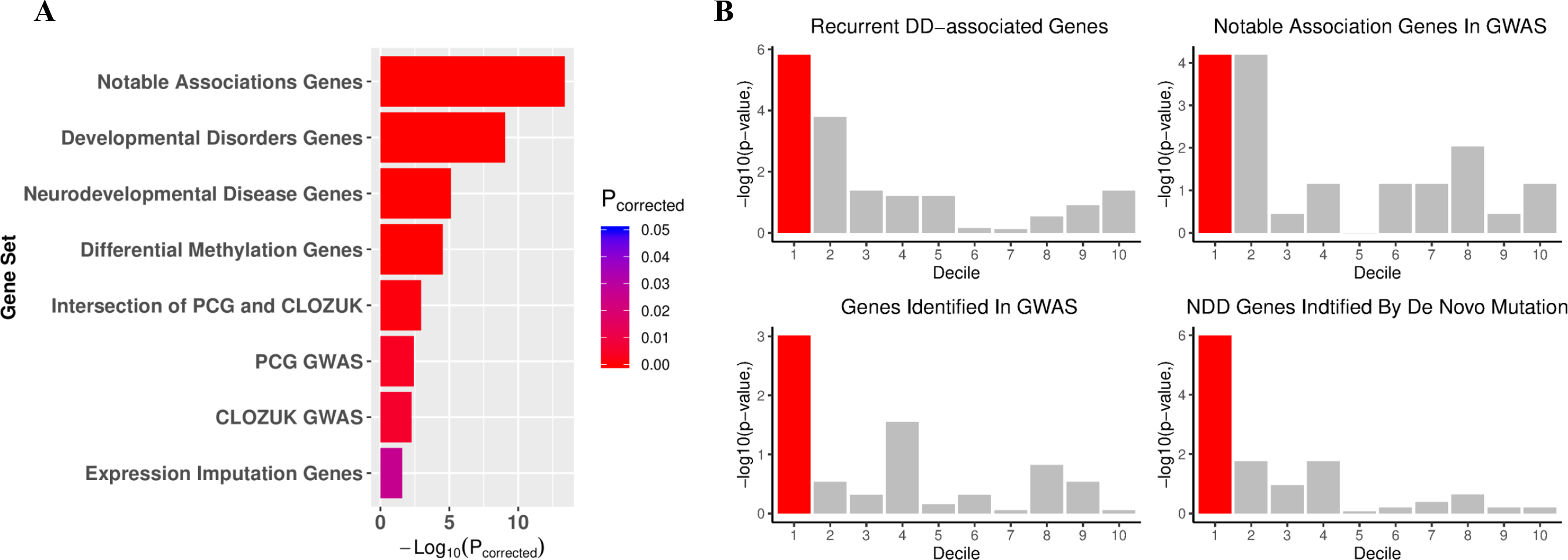
The evaluation of genes we predicted. (a) Validate the overlap of SCZ gene prediction within various gene sets from multiple genetic and epigenetic studies. The predicted gene shows significant enrichment in each SCZ-associated gene set (one-tailed Fisher’ s exact, FDR-adjusted). (b) Evaluation of the performance of our strategy on prioritizing the SCZ related gene, the first decile genes show the most significant enrichment (one-tailed Fisher’ s exact) compared with others.

These preliminary evaluations of our result indicated that our strategy acquired a reliable prediction of the SCZ-related genes. We found that the first decile gene set show more significant enrichment than genes sorted behind (Fig.2 B), and enrichment analysis based on DisGeNET data showed that schizophrenia is the most enriched disease in the first decile genes^16^, which indicate that the first decile genes can be regarded as high-confidence gene (HCG) for further analysis.

### Identifying causal genes of schizophrenia

While GWAS have identified over 100 loci associated with SCZ, only a few causal genes in these loci have been identified. Here we applied HMPDG for prioritizing genes in GWAS association loci. It is reported that the SCZ risk genes have more incoming regulatory links than background^17^. We observed that the PRGs in the GWAS loci indeed have more distal regulatory elements (DREs) in the germinal zone (GZ, P= 6.03 × 10^−3^) and, cortical and subcortical plate (CP, P=0.04) than Non-PRGs^18^. Neuroimaging studies have identified that the central nervous system and brain function of SCZ patients is significantly different from normal people. Therefore, we next explored gene-expression in the brain to test the hypothesis that SCZ-risk genes have a higher expression level in the brain. As expect, PRGs are highly expressed compared to Non-PRGs in the Allen brain data (P <2.2 × 10^−16^, Wilcoxon one-tailed test)^19^. Further, the PRGs are more intolerant of loss-of-function (LoF) mutation and under strong background selection (P =2.13 × 10^−4^, Wilcoxon one-tailed test)^20^.

In the largest CNV study to date, PGC CNV Analysis Group has reported eight genome-wide significant CNV loci, which involved hundreds of adjacent genes^21^. Yet, revealing the causal gene among a large number of genes disturbed by CNVs remains challenging. To meet this challenge, we applied HMPDG to systematically prioritize genes in the CNV region. We evaluate genes in seven widely reported SCZ-associated CNV loci (1q21.1,2p16.3,3q29,7q11.23,15q13.3,16p11.2(including distal 16p11.2 and proximal 16p11.2),22q11.2)^21^, and found that PRGs within these CNVs have more previous genetic or function evidence that supports them as important genes for SCZ(Fig.3 A). To investigate whether PGCs are more intolerant of haploinsufficient, we introduced the pLI-the probability of gene fall into haploinsufficient category-to assess the intolerance of PGCs and Non-PGCs affected by CNVs. We found that the pLI score of PRGs is higher than Non-PRGs (P =0.013, Wilcoxon one-tailed test) (Fig.3 B-1). As expected, the PRGs in the CNV region also have a higher expression level than Non-PRGs (P <2.2 × 10^−16^, Wilcoxon one-tailed test) (Fig.3 B-2). We outlined the biological processes enriched in the PRGs disrupted by CNVs, while no pathway enriched in the Non-PRGs. In Go terms significant for PRG, the majority consisted of synaptic, neuron, and protein kinase activity; in particular, the gene set related to synaptic is most abundant, which is consistent with the SCZ-related CNV studies^21^(Fig.3 C).

**Figure 3:**
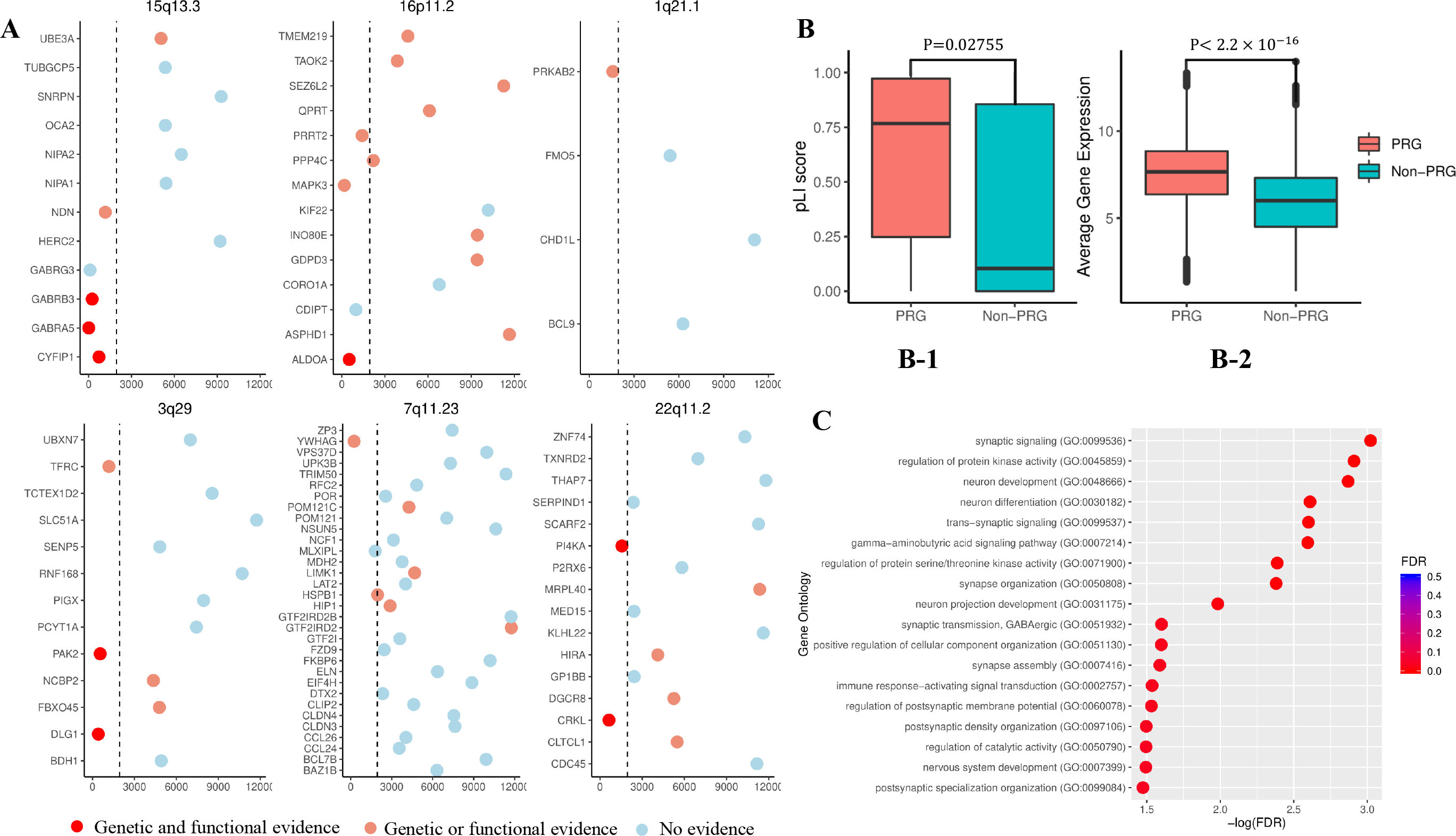
Reveal key schizophrenia genes affected by CNVs. (a) Each scatter plot represents the order of genes in the six CNV regions except 2p16.3 (only one gene in the 2p16.3), different colors indicate the degree of support of the evidence, the dotted line represents our decision boundary, the smaller the order(X-axis), the greater the probability of genes associated with schizophrenia. Compared to negative genes (non-PRGs) with bare evidence, most of the key SCZ-associated genes we predicted in the same CNV are supported by richer evidence. (b) PRGs have higher pLI score (b-1, p-value=0.02755, Wilcox test) RNA expression level in the brain (b-2, p-value<2.2 × 10^−16^, Wilcox test) and compared to Non-PRGs.(c)GO enrichment analysis of PRGs in the CNV region.

We reviewed the HCGs located in the GWASs loci or CNV region and, 12 promising causal genes were nominated by HMPDG (including *GRM3, GABRA5, DRD2, SLC32A1, CHRM4, GABRG3, RIMS1, STAT6, AKT3, MAPK3, MEF2C, GRIN2A*). Most of these causal genes are implicated in the hypotheses of etiology and treatment of SCZ, including glutamatergic neurotransmission(*GRM3, GRIN2A*), GABAergic synapse(*GABRA5, GABRG3, SLC32A1*), synaptic plasticity (*MEF2C*), neuronal calcium signaling (*RIMS1*) and the target of antipsychotic risperidone(*DRD2*) ^3^. Our analysis provided a well-founded list of the schizophrenia causal genes. Further exploration of these genes will facilitate insight into SCZ genetics and pathogenesis.

### Spatiotemporal expression pattern of schizophrenia risk genes

Although the knowledge of the genetic basis of schizophrenia has been improved recently, the link between these genes and the pathophysiological features of schizophrenia is not yet understood clearly. The emergence of single-cell RNA-sequencing can allow us to find the specific brain cell type underlying SCZ-related genes. Here, we employed the scRNA data from Karolinska Institute (KI) to investigate the specific-expressed cell type of SCZ by Expression Weighted Cell Type Enrichment (EWCE)^22-25^. We first evaluated the relation of HCG to the KI level 1 brain cell types. The HCG showed highly specific expression in four cell types: cortical interneurons, neocortical somatosensory pyramidal cells, striatal medium spiny neurons, and hippocampal CA1 pyramidal cells, which is almost identical to the study of Nathan G Skene et.al^24^.

The loss of heritability indicated that genes cannot achieve statistical significance remains to contribute to SCZ. We proposed that the specific-expression study of the full spectrum of SCZ genes may provide new insights into SCZ-related cell type. As expected, we found new cell types related to SCZ: Microglia cells, striatal interneuron cells, both of them have repeatedly implicated in SCZ^26-28^(Fig.4 A). Notably, the microglia cell plays a great role in the synaptic pruning, which has been hypothesized as the pathogenesis of schizophrenia^27^. However, previous cell type enrichment studies of SCZ missed association of SCZ gene set with microglia^24,29^. Our result provided the evidence for the hypothesis that microglia may etiologically involve in SCZ.

**Figure 4:**
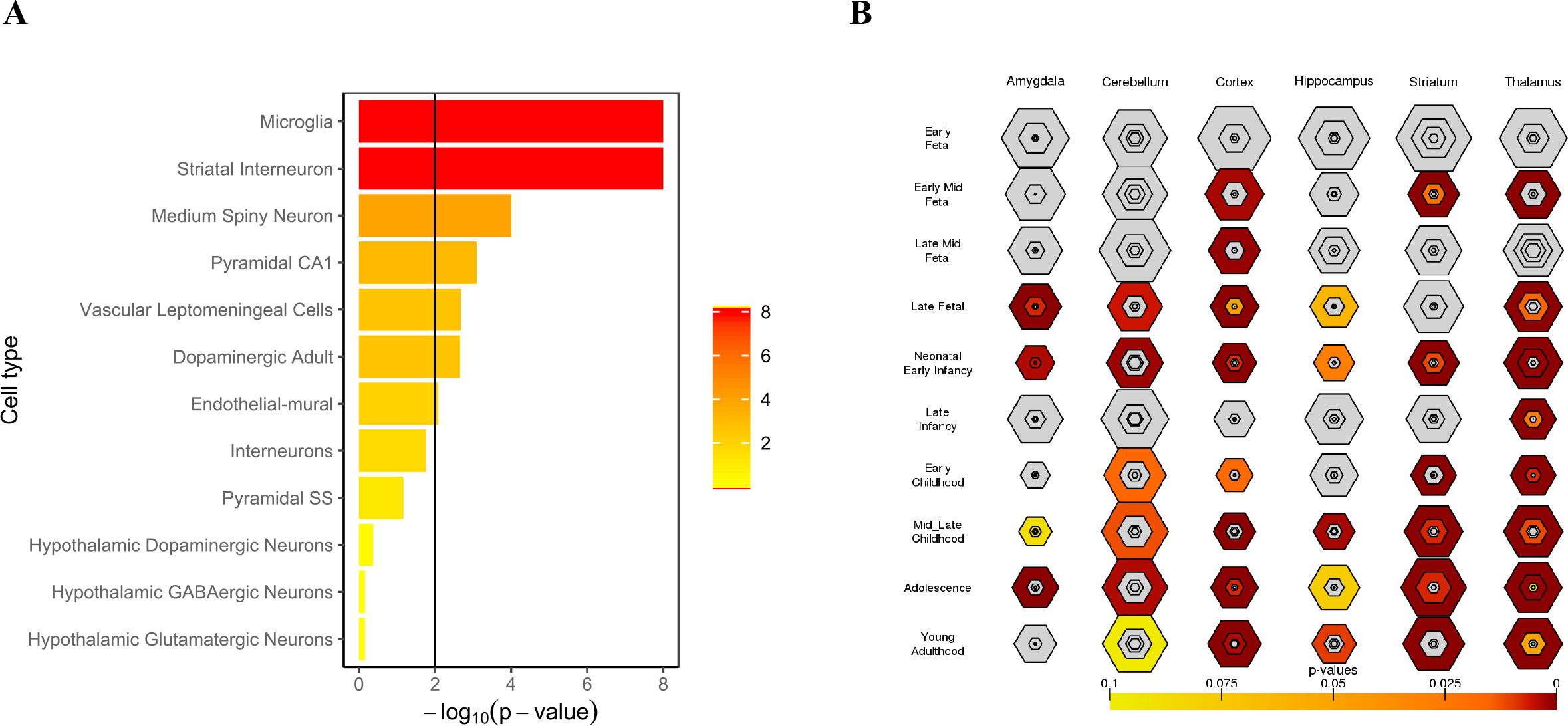
(A) evaluation of enrichment of SCZ genes in scRNA-seq data from KI (B)developmental expression pattern of SCZ-associated genes we predicted. The size of the hexagon corresponds to the number of genes and the color shows the significance of the P values (one-tailed Fisher’ s exact, FDR-adjusted). (A) Cell type-specific expression analysis showed that microglia cells, striatal interneuron cells, striatal medium spiny neurons, and hippocampal CA1 pyramidal cells play critical roles in SCZ pathophysiology. (B) Temporal expression analysis of PRG highlights a strong signal of various parts of the developing brain in late fetal to early infancy and puberty.

Based on the BrainSpan database, we performed specific enrichment analyses across brain regions and development stages for candidate genes^30,31^. PRGs are significantly enriched across many brain regions and different stages of development. Among these, we observe the developmental pattern of SCZ-risk genes both highlight in the prenatal and adolescence. This pattern is in accord with previous studies on the development stages of SCZ^32^, especially with recent findings that adolescence is a critical period for SCZ^33^(Fig.4 B). Spatially, we find that SCZ risk genes widely affect various regions of the developing brain, which illustrated the underlying mechanism of phenotypic heterogeneity in schizophrenia^34^. Extensive literature implicates cortex, striatum, and thalamus in the pathophysiology of this disorder^35-37^, and our analysis shows that these regions in neonatal early infancy and adolescence get the most profound enrichment(Fig.4 B). This finding demonstrated that the disrupted function of cortex, striatum, and thalamus at the neonatal early infancy and adolescence may contribute to SCZ risk. In addition to the known dysfunction region in SCZ, the amygdala was illustrated as a critical locus of schizophrenia pathology in adolescence, a finding of relevance to reducing amygdala volume in early course schizophrenia^38^.

### Functional modules disrupted in schizophrenia

Recent genetic and experimental studies on SCZ have discovered a variety of cellular functions and pathways disrupted in patients such as dopamine model ^39^ and glutamate model^40^, which proposed diverse hypotheses for the molecular basis of the etiology of SCZ. Figuring out the effect of the SCZ related genetic changes in biological function will result in a comprehensive understanding of the molecular mechanism of SCZ. Since the known schizophrenia gene is incomplete, however, it cannot provide a global schizophrenia functional map. Here, our gene full spectrum prediction results make it possible to deconvolve SCZ-related genes into detailed biological pathways. We performed MCODE, a graph-theoretic clustering algorithm^41^, to isolate the dense sub-network in PPIN of the PRG. Then we made functional annotations for each group by Go analysis. Finally, we shaped a complete functional map of SCZ with different and substantially cohesive functional modules affected by PRGs (Fig.5).

**Figure 5:**
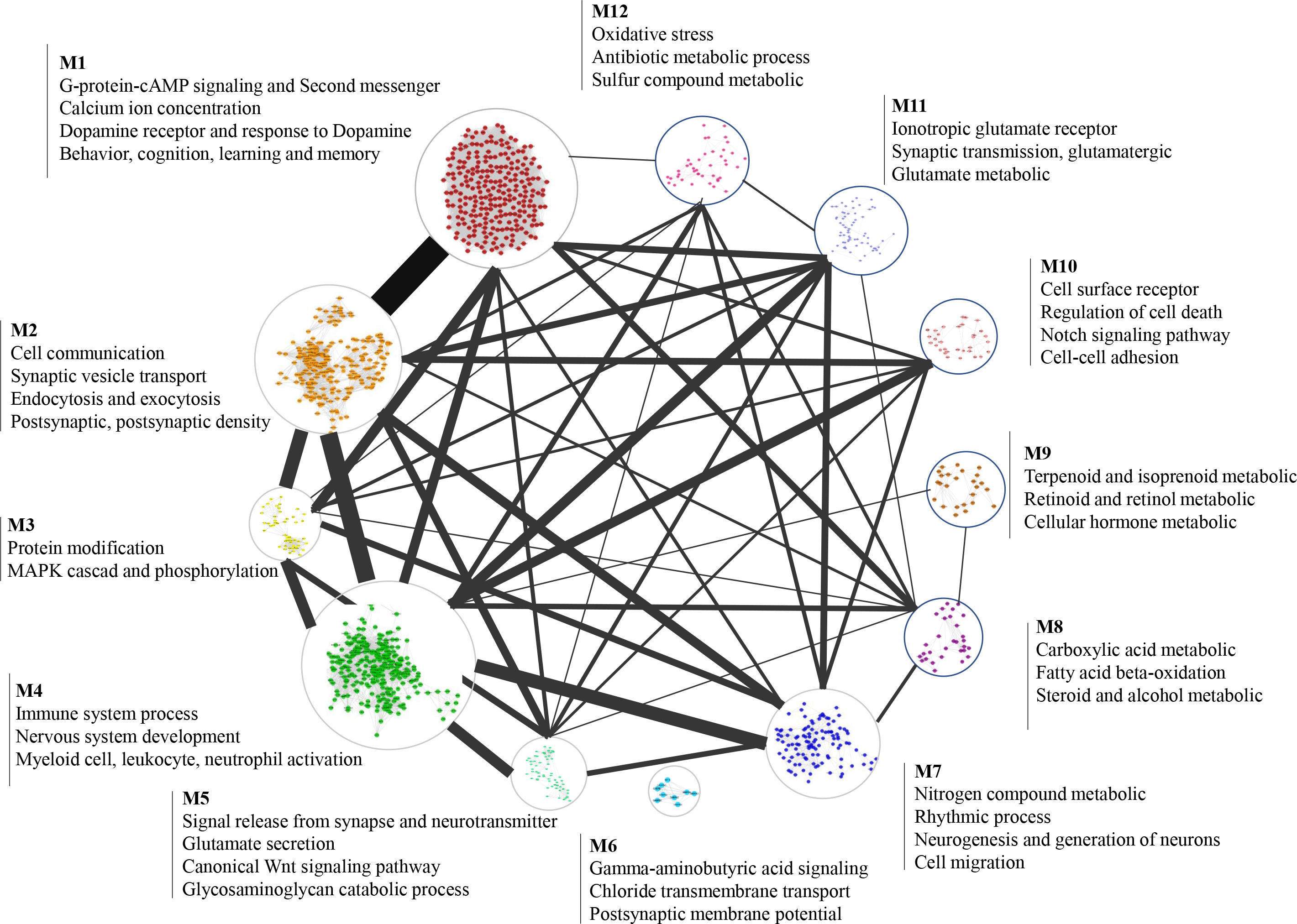
Graphical sketch of the 12 most populated functional modules of the SCZ-associated genes. Each module is linked by lines based on PPI evidence from STRING and, the width of the line consists of the number of links. The size of each module corresponds to the number of genes and the notes are screened from GO terms with the most significant P-value (one-tailed Fisher’ s exact, FDR-adjusted).

This strategy provides a systematic method for exploring the impaired molecular function and the molecular basis of SCZ. We have observed several functional modules are consistent with the classic etiology hypothesis of schizophrenia, including dopamine hypothesis^39^(dopamine receptor and response to dopamine, classified as module 1(M1)), glutamate hypothesis^40^ (ionotropic glutamate receptor-N-methyl-D-aspartate receptor(M11)), neurodevelopment hypothesis^42^ (this annotation exist in multiple modules, such as M3 and M7). Furthermore, our systematic analysis results advocate retinoid as a strong contender to the new hypothesis of mechanisms of SCZ, which have multi-lines of evidence support that dysregulation of the retinoid system involved in the etiology of SCZ^43^. Modules we depict here also link genes with phenotypes of the SCZ patients. For instance, genes implicated in cognition, learning, and memory (M1) might underlie the cognitive deficits that earliest and constantly found in schizophrenia patients. Similarly, patients with SCZ frequently suffer insomnia and disrupted sleep-wake cycles^44^, and the genes clustered in C7 are involved in circadian rhythm. More interestingly, we recognized some functional modules of schizophrenia that are non-classic etiology hypothesis of SCZ and received little attention previously. The oxidative stress that captured in M12 is a good illustration, genes in M12 are involved in the synthesis of glutathione, peroxiredoxin, and superoxide dismutase, which have reported contribute to the pathophysiology of SCZ in previously studies^45^. Also, noteworthy functions of glycosaminoglycan that identified in M5 including axon guidance, neuronal migration, regulation of synaptic, and neurotransmission are highly in line with the pathophysiology of SCZ^46^, which implies the glycosaminoglycan abnormality is a potential candidate for the etiology of SCZ.

These findings reveal the functional role of PRGs and provide a global perspective for better exploring the mechanism underlying the phenotype and pathophysiology of schizophrenia. We carefully examined all the functional roles of PRGs involved (including those not shown in the figure such as cardiac conduction, autophagy, and inositol phosphates), and most of these functional modules have varying degrees of evidence to support these modules related to the pathogenesis of SCZ. Further, our findings have suggested some potential biomarkers of schizophrenia with evidence but not adequate from previous studies^47^, such as glutamate (M5), Gamma-aminobutyric acid (M6), and arachidonic acid(M14) and we emphasize that further study is needed to validate these metabolites.

## Discussion

Schizophrenia is a complex mental illness characterized by multiple modes of inheritance and daunting etiology. Exploring full-scale risk genes, in our perspectives, is the first step to elucidate the genetic and pathophysiology of schizophrenia. Yet, it is unlikely to discover the full spectrum of risk genes based sequencing studies alone. Here we put forward an integrative predict strategy based on systems biology, HMPDG, to provide a complement of SCZ risk genes. The validity of risk genes we predicted was attested to by multiple independent genetic studies, demonstrating the promise of these genes to gain new insights into SCZ genetics and pathogenesis. Since this strategy has excellent performance and repeatability, we believe that it can be promoted to other complex disease studies.

Identifying causal genes in sequencing-based associations is a critical step to bridge genes and biology of schizophrenia. Underlying the comprehensive risk gene spectrum, we provided a high confidence SCZ-related gene list and revealed causal genes among various variants in associated loci and CNV regions of SCZ. The causal genes in the CNV region and GWAS show high intolerance to haploinsufficient. This character corroborated with the hypothesis that SCZ-related genes are under strong background selection and indicated that the causal genes are more likely to cause functional impairment than non-PRGs in GWAS loci and CNV region^6^. What’s more, the causal genes have higher expression levels and more DREs in the brain than no-PRGs, suggesting that these genes may play a critical role in the central nervous system dysfunction of SCZ. Taken together, we provide a reliable causal gene list for SCZ; furthermore, the strategy we performed demonstrated how researchers can use the biological network and gene pathogenicity score that is prevalent in prioritizing Mendelian disease candidate genes to identify the causal gene for complex disorders.

Depicting the full spectrum of risk genes and pinpointing causal genes for schizophrenia are not the finale of our study, we also performed empirical research based on predicted complete component for SCZ genes for revealing comprehensive insight of schizophrenia etiology. We reconfirmed brain cell types underlying schizophrenia identified in the previous study and revealed two additions: microglia and striatal interneuron that are also prominent in the etiology of schizophrenia^24^. It is reported that striatal interneurons are involved in the pathophysiology of SCZ. As the resident macrophage cell, microglia play a great role in excessive synaptic pruning, which contributes to the reduction in synapse density in SCZ patients^27^. Although various studies have identified that the microglia indeed involved in the etiology of SCZ, recent studies that focus on cell type implicated with SCZ are not consistent with these findings^24,29^. Here, we reported that the genes implicated in SCZ mapped onto microglia and striatal interneuron for the first time. Furthermore, we reveal that these genes specific to microglia are relevant to complement signaling and immune response, including *C5AR1* and *C3AR1* which have reported to associated with SCZ in functional experiments^48,49^. Enrichment analysis of the top 100 genes specific to microglia showed that a significant signal in the late fetal amygdala. Early developmental damage to the amygdala may result in ‘theory of mind’ (ToM) impairments, which are considered as core neurocognitive deficits in schizophrenia^49,50^. Here we emphasize that the potential role of the amygdala in schizophrenia should not be neglected.

In conclusion, our strategy provided a paradigm for combining genetic pathogenicity assessment with biological networks and specific RNA expression data to predict genes related to complex diseases. The full spectrum of SCZ risk genes showed the landscape of the genetic basis of schizophrenia. Of these, causal genes with genetic evidence represent a starting point for further mechanism study. Strikingly, exploratory analysis of these genes revealed new insight on the pathophysiology of SCZ. We expect that these genes and related biological processes, as well as associated spatiotemporal context, will further enhance the understanding of the genetic and etiology basis of schizophrenia and provide help for the diagnosis and treatment of schizophrenia.

## Methods

### Data for build model

The protein-protein interactions (PPI) data were download from the STRING v11^51^ database. To reduce noise, we selected PPI with the highest confidence score (≥0.900). Finally, we obtain 505,154 PPIs between 12,271 proteins and then constructed a relatively objective and comprehensive protein interaction network. Neurodevelopmental RNA-seq data were obtained from the BrainSpan Atlas^52^. The expression value of each gene was calculated by the average of all samples. GeVIR^53^, which outperforms missense constraint metrics, was introduced to provide pathogenicity score for each gene. According to the assessment by Xiang Yue1 et al.^54^, compared with other embedding methods, node2vec performs well in node classification tasks based on PPIN^55^. So as to preserve the homophily of each node in the PPIN, through a hyperparameter search, the settings of p = 4 and q = 1 were used here. Finally, we obtained a 128-dimensional vector representation of each gene in our PPIN via the node2vec algorithm. Based on previous evaluations of centrality measurements for the detection of essential genes or protein in biological networks, we selected 13 topological parameters to identify those genes that play a major role in the pathogenic mechanism or functional activities^56,57^. The entire table of these parameters is provided in Supplementary Table 1.

### Predicting Genes associated with schizophrenia

Random Forest Classifier was recruited to deal with this classification task. We used the random forest to build the prediction model and adjusted the parameter by the five-fold cross-validation. Features used for classification include embedded low-dimensional vectors(n=128), 13 topological parameters, RNA expression values(n=1), and gene pathogenicity scores(n=1). We curated 218 genes linked with SCZ from SCZ-related gene dataset^58^, 187 genes were finally enrolled due to incomplete PPIN data (12,271 proteins). Genes annotated to non-diseases and non-essential as negative examples^59^, after filter by Tajima’s D test,291 genes were employed.

### Gene set enrichment analysis

We collated the genes corresponding to loci reported in the previous GWAS studies and matched them with our data^3,6^, (keep the genes in our data set and remove the genes in the training set), then we get 4 SCZ-associated gene sets: 1) associated gene set from PGC,2)notable association gene set from PGC,3) associated gene set from CLOZUK and 4) intersection of associated gene sets from PGC and CLOZUK.

Considering the transcriptomic studies play a great role in complex disease studies, we then select a schizophrenia gene set provided by gene expression imputation study across multiple brain regions to check the enrichment of our predict genes^13^. What’s more, we collected SCZ related genes from 6 methylation studies included in SZDB2.0^7-12,60^, which contains 3 brain prefrontal cortex researches and 3 blood researches. In order to minimize the impact by the false positive and confounding factors such as age, gender, and experimental batch, we selected the recurrent genes that detected in no less than 2 studies as SCZ related risk methylation loci. In addition to the well-established SCZ genes, we also collected the neurodevelopmental disease gene set and the developmental disorders gene set from DECIPHER to explore the genetic overlap between SCZ and development disorders.

One-sided Fisher’s exact tests were adopted to evaluate the enrichment of our prediction with known SCZ-associated genes and multiple tests were corrected by Benjamini–Hochberg corrections.

### Cell type enrichment analysis

One recent study reported the brain cell type of SCZ base on the single-cell RNA data of KI. The cell-type expression specific matrix of KI was download (http://www.hjerling-leffler-lab.org/data/scz_singlecell/). The EWCE package was adopted to find in which brain cell type the predict SCZ-associated genes have higher expression levels. The gene set enrichment analysis across brain regions and development was performed by Cell-Specific Expression Analysis (CSEA) tool^30,31^.

### Identifying functional modules

The PPIN of PRGs were obtained from the STRING v11 database. To cluster the network, we used MCODE to find tightly connected sub-network. Then we performed GO analysis for each sub-network to find the GO biological processes of these genes in each sub-network. The modules have a similar function have divided into one group and, the modules with few genes are not shown in the figure. The detail of modules and these functions are provided in Supplementary Table 4.

## Data Availability

This study makes use of data generated by the DECIPHER community. A full list of centres who contributed to the generation of the data is available from https://decipher.sanger.ac.uk and via email from. Funding for the DECIPHER project was provided by Wellcome

## Author contributions

JF.L., JG.Z, and HK.L. conceived the study. JF.L.L.W. WM.F., YT.M, and L.L. collected the data. JF.L., SJ.H. and ZW.W. performed the *in-silico* analysis. JG.Z. supervised the research. JF.L. wrote the first draft of the manuscript. HK.L., J.W., and JG.Z. reviewed the manuscript. All authors revised the manuscript. All authors had full access to the final version of the manuscript and agreed to the submission.

## Acknowledgments

This study was supported by Shenzhen Municipal of Government of China (NO. JCYJ20170412153248372), Shenzhen Municipal of Government of China (NO. JCYJ20180507183615145), The National Key R&D Program of China (NO.2016YFC1305900), The National Key R&D Program of China (NO.2017YFC1308402), The National Natural Science Foundation of China (NO.81771444).

